# Prevalence of Shigellosis in Pediatric Diarrheal Patients in Chattogram, Bangladesh: A Molecular Based Approach

**DOI:** 10.1101/2022.09.16.22280035

**Authors:** A K M Zakir Hossain, Md. Zahid Hasan, Sohana Akter Mina, Nahid Sultana, A M Masudul Azad Chowdhury

**Author notes:** These authors contributed equally to this work (A K M Zakir Hossain, A M Masudul Azad Chowdhury).

## Abstract

*Shigella* a gram negative, non-motile bacillus, is the primary causative agent of the infectious disease shigellosis, which kills 1.1 million people worldwide every year. The children under the age of five are primarily the victims of this disease. This study has been conducted to assess the prevalence of shigellosis through selective plating, biochemical test and conventional PCR assays, where the samples were collected from suspected diarrheoal patients. Invasive plasmid antigen H (*ipaH*) and O-antigenic *rfc* gene were used to identify *Shigella spp*. and *S. flexneri* respectively. For validation of these identification, PCR product of *ipaH* gene of a sample (Shigella flexneri MZS 191) has been sequenced and submitted to NCBI database (GenBank accession no-MW774908.1). Further this strain has been used as positive control. Out of 204, around 14.2% (n=29) pediatric diarrheoal cases were screened as shigellosis. Another interesting finding was that most of shigellosis affected children were 7 months to 1 year. The significance of this study lies in the analyses of the prevalence and the molecular identification of *Shigella spp*. and *S. flexneri* that can be utilized in improving the accurate identification and the treatment of the most severe and alarming shigellosis.

## Introduction

The diarrhea is the major symptom of shigellosis which is an acute intestinal infection [1] caused by gram-negative, facultative, enteric intracellular pathogenic bacteria *Shigella*. Shigellosis ranges from mild watery diarrhea to severe bacillary dysentery with fever, abdominal pain, and frequent passage of bloody, mucoid and small-volume stools [2] [3]. High fever, strong abdominal cramps and a mucopurulent diarrhea with blood are the symptoms of shigellosis [4,5]. Shigellosis leads to death and constitutes one of the major morbidities and mortalities among infant and children worldwide [6]. It was estimated that worldwide *Shigella* causes 164.7 million cases of shigellosis with 163.2 million from developing countries resulting 1.1 million deaths annually [7]. Recently in Asia, it was reported that the number of shigellosis cases were at nearly 91 million, resulting in 414,000 deaths each year [8]. There is no exact clear demography, however, some previous studies conducted in six-Asian countries (Bangladesh, Pakistan, Thailand, China, Vietnam and Indonesia), the burden of *Shigella* infection rates were high in children under the age of 5 [9].

There are four species of *Shigella* based on antigenic properties: *S. dysenteriae, S. flexneri, S. boydii and S. sonnei* [10]. Among these, *Shigella flexneri* is mostly isolated in developing countries [5]. In Bangladesh, *S. flexneri* is the most frequently isolated (54%-60%) gastrointestinal pathogen [11] and it has been estimated that annually more than 95,000 children<5years of age die due to shigellosis [3] [12]. *Shigella flexneri* is highly infectious and 10^2^ -10^3^ bacteria is suffice to cause diarrhea in human [13][14]. This low infective dose has ability to survive in low acidic environment in human stomach by up regulating the acid resistance genes [15]. *Shigella flexneri* invades the colonic and rectal epithelium of primates and humans not through apical surface, it uses the antigen sampling microfold cells (M cells) and basolateral surface of its target cells, causing the acute mucosal inflammation characteristic of shigellosis [16]. Now shigellosis is a major public health concern in developing countries like Bangladesh where people live in poor sanitation and in overcrowded condition [14].

The epidemiological data of Shigellosis in Bangladesh perspective and the proper screening of *Shigella spp* as well as *Shigella flexneri* are needed to conduct a proper treatment. The effector invasive plasmid antigen H (*ipaH*) gene and other invasive plasmid antigens (*ipa*) can be used as the potential target for molecular identification of *Shigella* [1]. The IpaH is a unique polypeptide that is found only in *Shigella* and *Enteroinvasive Escherichia coli* (*EIEC*) [1]. On the other hand, the O-antigenic *rfc* gene is excellent candidate for *S. flexneri* identification among *Shigella spps*. as this gene is specific for *S. flexneri* [5]. We conducted this study to confirm a rapid, accurate and a reliable method for identification of *Shigella* and *S. flexneri*.

After accurate identification, we show the prevalence of Shigellosis caused by *Shigella spp*. as well as *Shigella flexneri* in the case of pediatric diarrheoal patients in Chattogram, Bangladesh.

## Material and Methods

### Sample Collection

To conduct research with human clinical samples, we first obtained ethical approval from the “Institutional Review Board” of Chattogram Maa-O-Shishu Hospital Medical College, Chattogram, Bangladesh (Ref: CMOSHMC/IRB/2020/03). Stool samples were collected from the diarrheal word, Chittagong Maa-O-Shishu Hospital Medical College and stored at -20ºC in our ‘Laboratory of Microbial and Cancer Genomics’, Department of Genetic Engineering and Biotechnology, University of Chittagong, Chattogram, Bangladesh. **Sample Culture:** The samples were cultured on XLD agar (Xylose Lysine Deoxycholate) medium for primary screening. Then the pink colonies were transferred on MacConkey agar for pure culture and transparent colonies were suspected for *Shigella flexneri*.

### Biochemical test

The triple sugar iron (TSI) agar medium was used for biochemical test. The inoculated TSI test tubes were incubated at 37°C for overnight. The TSI test tubes within alkaline slant and acidic butt without gas and H_2_ S were suspected for *Shigella spp*. and *S. flexneri*. These suspected samples were selected for DNA isolation and molecular identification. Other TSI test tubes (alkaline slant + acidic butt with gas or H_2_ S, alkaline slant + alkaline butt, acidic slant + acidic butt with and without gas or crack) were rejected.

### Suspected Sample Storage

The nutrient agar slant was used for short-term storage and 10% skim milk was used for long-term storage.

### DNA extraction

This study used the boiling method to isolate bacterial DNA [17]. After purity and DNA concentration check through NanoDrop, highly purified and concentrated DNA samples were selected for PCR amplification and stored at -20ºC.

### Molecular identification of *Shigella spp*. and *S. flexneri*

The primer pairs of *ipaH* and primer pairs of *rfc* gene (Table-1) were designed. PCRs were performed in a 10 µl mixture, composed of 5 µl of PCR master mix (Promega), 0.5 µl forward, 0.5 µl reverse primers (10 mmol/ml), 3 µl nuclease free water and 1 µl of sample DNA template. For *ipaH* gene identification, PCR amplification reactions were performed in a PCR machine (Nyx Technik, Inc. model: A6, USA) under the following conditions: 95 °C for 15 min and then 33 cycles of 15 S at 95 °C, 30 S at 58 °C, 30 S at 70 °C and one cycle final elongation was performed for 7 min at 72 °C. For *rfc* gene identification, PCR amplification conditions were 95 °C for 3 min and then 35 cycles of 35 S at 95 °C, 35 S at 48 °C, 25 S at 72 °C and one cycle final elongation was performed for 2 min at 72 °C. Amplified PCR products were visualized and verified by separating and comparing the DNA bands with appropriate DNA ladder and positive control (*Shigella flexneri*_MZS_191, GenBank accession no: MW774908.1) through electrophoresis in 1.5% agarose gels with ethidium bromide.

**Table 1:**
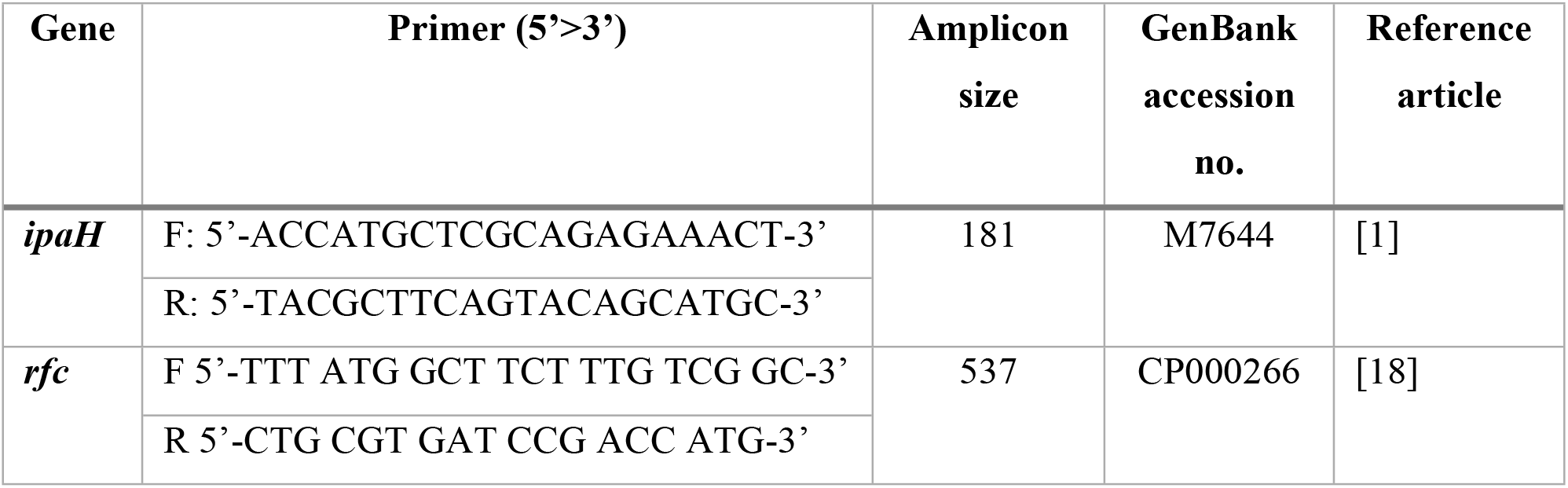

### Statistical Analysis

Statistical analysis were done by SPSS.

## Result

### Isolation and identification of *Shigella spp*. through selective plating and TSI

The characteristic pink colonies (Fig 1A) on XLD (Xylose Lysine Deoxycholate) medium were suspected for *Shigella* and *S. flexneri*. Through direct stool culture on the XLD agar plate, 107 samples out of 204 showed pink colonies. In pure culture the pink colonies formed transparent colonies (Fig 1B) on MacConkey agar plate. These transparent pure colonies were used for biochemical test TSI. Red/acidic slant and yellow/basic butt indicated *Shigella* (n=29) (Fig 1C). By selective platting, and biochemical test-TSI, 71 *Salmonella paratyphi*, 2 *Salmonella typhi* and 5 *Pseudomonas aeruginosa* were suspected.

**Fig 1:**
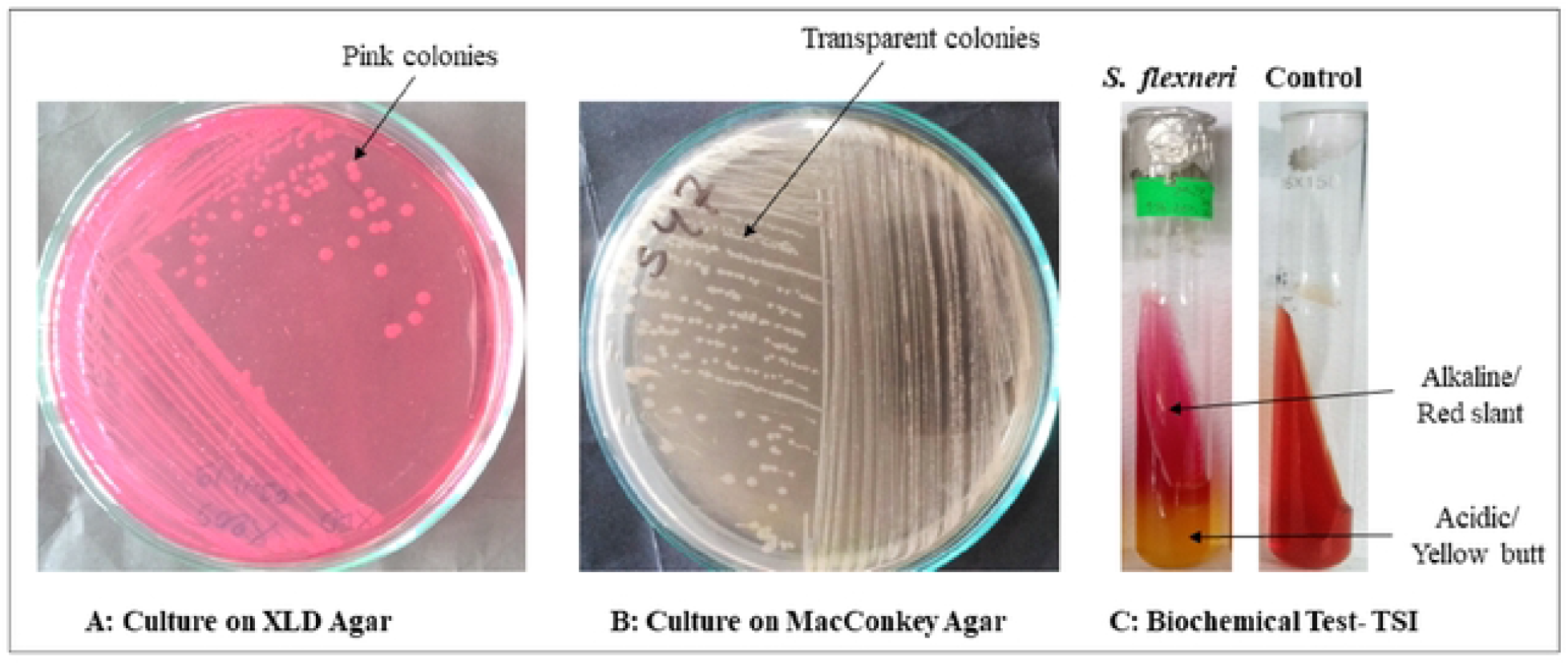
Selective plating and Biochemical Test. A: Target bacterial screening through XLD agar medium. Pink colonies were suspected as *Shigella*. B: Selected pink colonies were cultured on MacConkey agar medium and transparent colonies were selected for biochemical test TSI. C: On TSI biochemical test, acidic slants and alkaline butts without bubble were suspected as *Shigella spp*. and *Shigella flexneri*.

### Identification of *Shigella spp*. through *ipaH* gene amplification

The *ipaH* gene was identified in all suspected *Shigella* isolates. *S. flexneri* strain MZS-191 (GenBank Accession No. MW774908.1) was used as positive control (Fig 2A).

**Fig 2:**
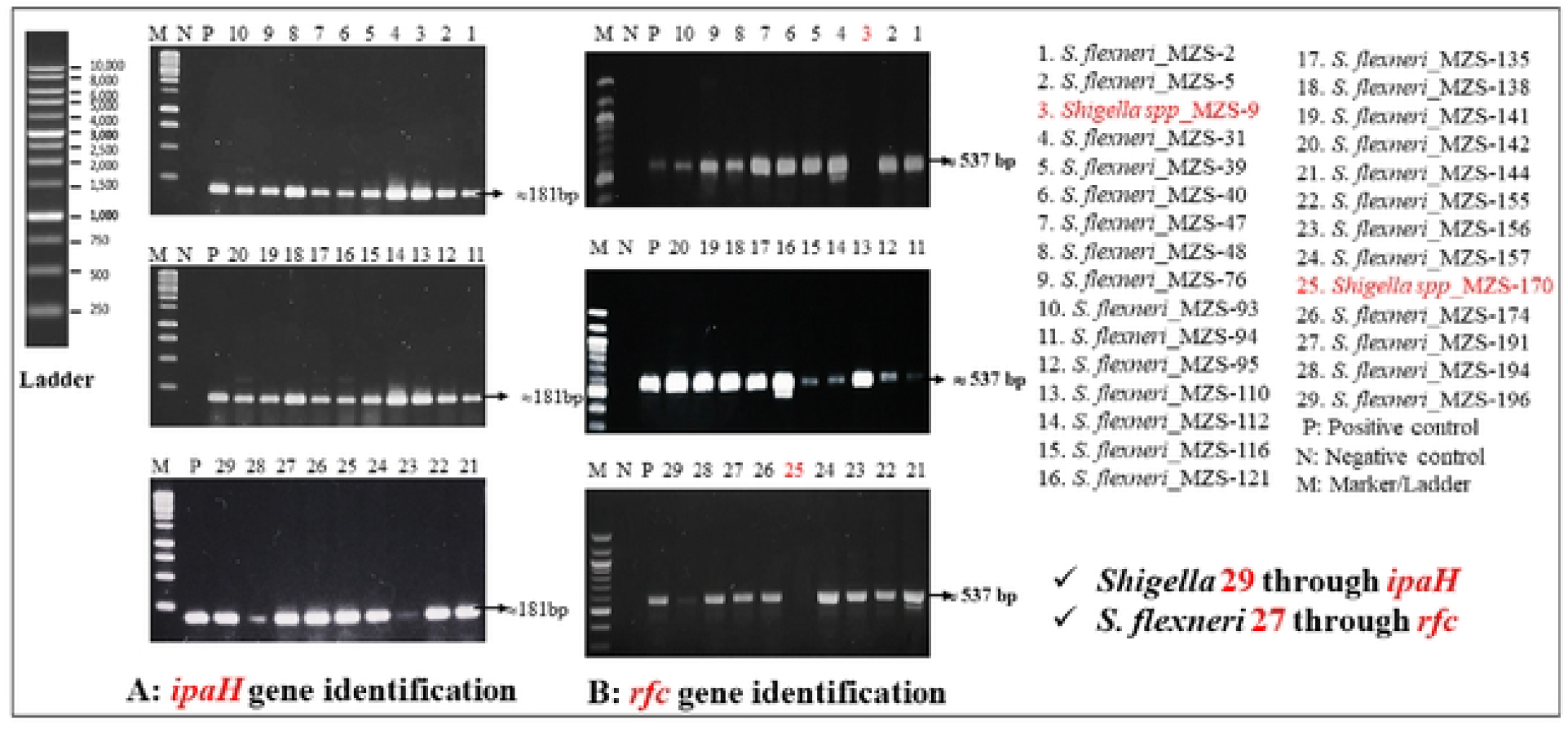
Molecular Identification of *Shigella spp*. and *Shigella flexneri:* Identification of the invasive *ipaH* and O-antigenic *rfc* gene in suspected gene in *Shigella flexneri* and *Shigella spp*. The target *ipaH* and *rfc* gene were amplified through PCR by using specific primers. After the completion of PCR, the PCR products were visualized through the agarose gel electrophoresis experiments. In both cases, *S. flexneri* MZS-191 (GenBank Accession No. MW774908. 1) was used as positive control. A: The *ipaH* was found in all suspected samples and confirmed that all of the 29 suspected samples were *Shigella flexneri* or *Shigella spp*. B: Since the *rfc* gene is *S*. *flexneri* specific, it was only found in *S*. *flexneri*. The *rfc* gene was identified in 27 samples among 29 *ipaH* containing samples. These 27 samples were confirmed as *Shigella flexneri*. The *rfc* gene was not identified in samples MZS-9 and MZS-25, which confirmed that the two were other species of *Shigella*.

### Identification of *Shigella flexneri* through *rfc* gene amplification

This *rfc* gene was detected in 27 samples among 29 suspected *Shigella flexneri. S. flexneri* strain MZS-191 (GenBank Accession No. MW774908.1) was used as positive control (Fig 2B).

### Prevalence of diarrheoal infections and Shigellosis

We obtained some most interesting data through selective plating, biochemical tests and molecular identifications. Different organisms were identified in diarrheoal pediatric patients (Table-2). We found that 14.2% (Fig 3A) diarrheal case was shigellosis and boys were more likely to be infected than girls (Fig 3B). In shigellosis cases, 93% (n=27) were caused by *S. flexneri* and 7% (n=2) were caused by other *Shigella spp*. (Fig 3C). Our findings also showed that all of shigellosis patients except one were less than five years old. In this rang of age intervals, the maximum infant (n=17) were 7 month to one years old. Three infants of shigellosis were 1 day to 6 months (Fig 4B). Not only Shigellosis, other diarrheoal pediatric patients mostly were in 7 months to 1 year of age rang (4B).

**Table 2:** Prevalence *of E*.*coli/ Klebsiella, Pseudomonas aeruginosa, Salmonella paratyphi, Salmonella typhi, Shigella spp, and Shigella flexneri* among diarrheic male and female children under-5 years of age in Chattogram, Bangladesh.

| Variables |  | Frequency | Percent (%) | Sex |  | P value |
| --- | --- | --- | --- | --- | --- | --- |
|  |  |  |  | Male | Female |  |
| Microorganisms | *E.coli/ Klebsiella* | 97 | 47.5 | 37 | 60 | 0.592 |
|  | *Pseudomonas aeruginosa* | 5 | 2.5 | 1 | 4 |  |
|  | *Salmonella paratyphi* | 71 | 34.8 | 27 | 44 |  |
|  | *Salmonella typhi* | 2 | 1.0 | 0 | 2 |  |
|  | *Shigella spp* | 2 | 1 | 0 | 2 |  |
|  | *Shigella flexneri* | 27 | 13.2 | 12 | 15 |  |
| Shigellosis | Negative | 175 | 85.8 | 110 | 65 | 0.663 |
|  | Positive | 29 | 14.2 | 12 | 17 | 0.405 |
| Age | 1D ≤ 6M | 32 | 15.7 | 20 | 12 | 0.199 |
|  | 7M ≤ 1Y | 94 | 46.1 | 59 | 35 |  |
|  | 1Y 1M ≤ 5Y | 72 | 35.3 | 45 | 27 |  |
|  | 5Y < | 6 | 2.9 | 3 | 3 |  |

\*p values were calculated by SPSS data analysis tool.

**Fig 3:**
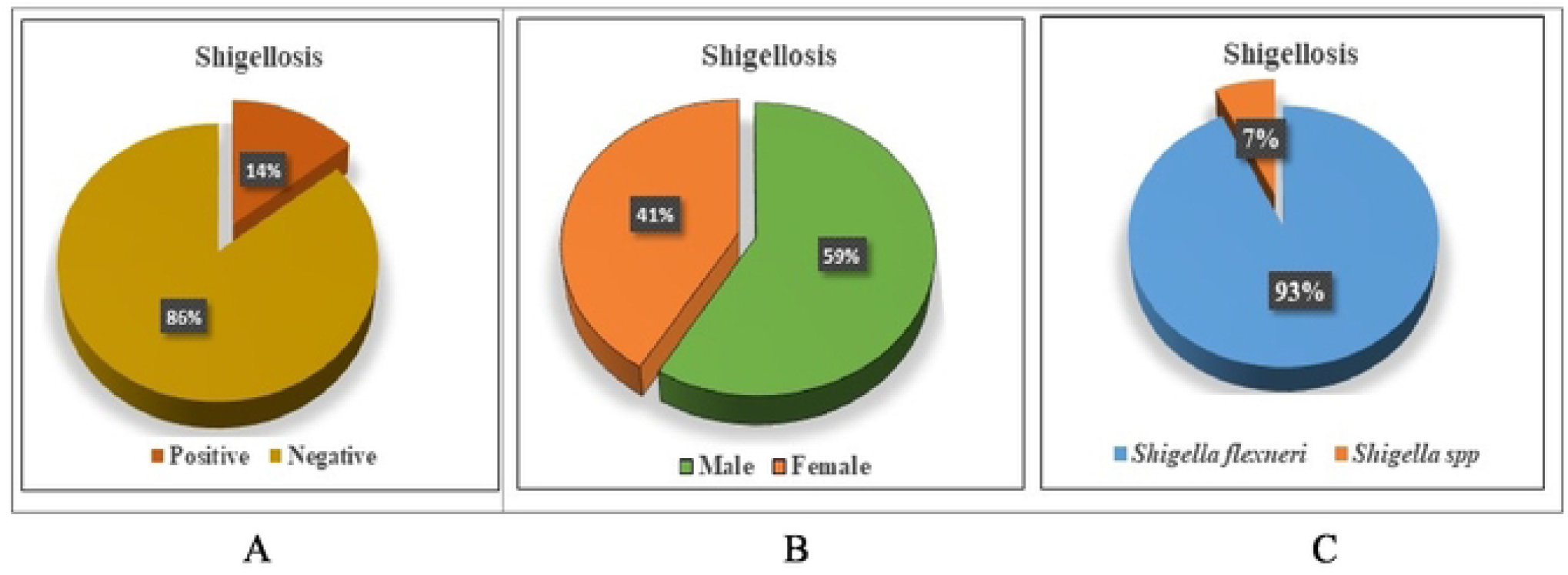
Shigellosis in different parameter. A: Among 204 diarrheoal patients, 14.2% (n=29) cases were Shigellosis. B: Most of patients were male children and C: among Shigellosis children, 93% (n=27) were infected by *Shigella flexneri*.

**Fig 4:**
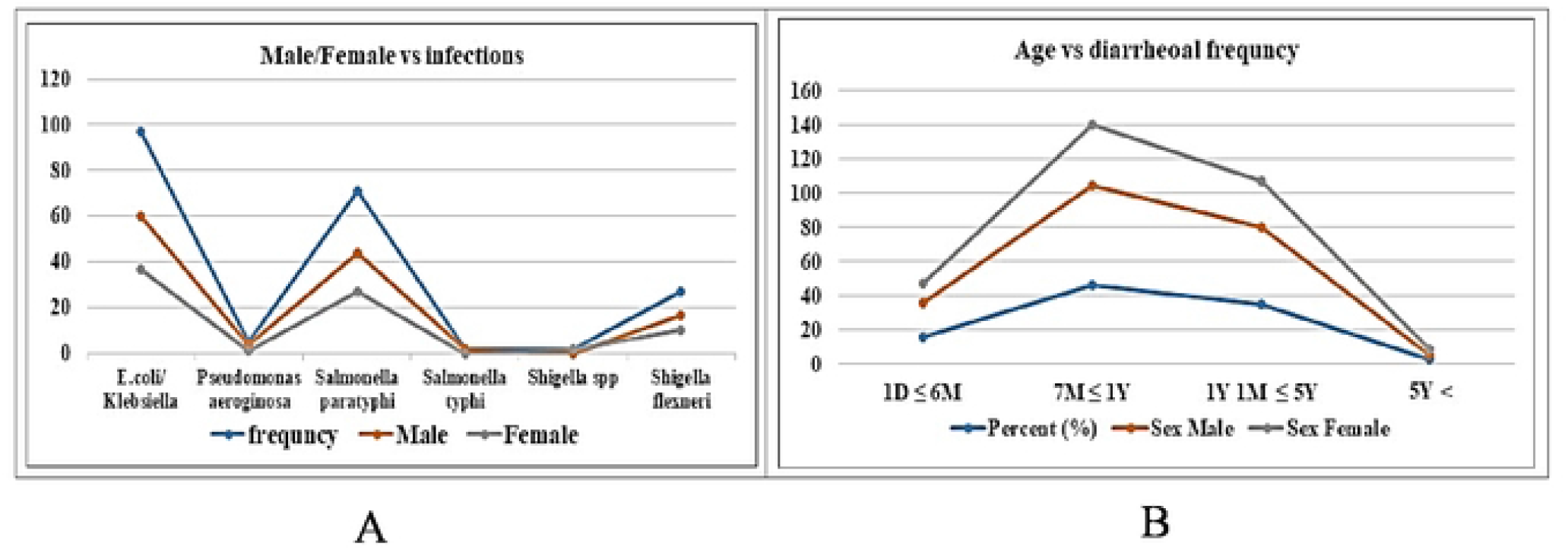
Frequency of organisms’ occurrence in diarrheoal patients regarding male and female children under different age interval. A: The graph represents that among 204 diarrheoal children most of the identified organisms were *E. coli/Klebsiellla* (n=97), *Salmonella paratyphi* (n=71), *Shigella flexneri* (n=27), *Pseudomonas aeruginosa* (n=5), *Shigella spp* (n=2) and *Salmonella typhi* (n=2). All of these bacteria are pathogenic and cause diarrheoal diseases in the host. Male children were found to be more infected than female children. B: Most of infected children were 7 month to 1 year interval.

## Discussion

Shigellosis, the most communicable bacterial diarrhea, occurs as a sporadic case. [18]. In low-income countries, this alarming disease sometimes outbreaks and causes epidemics and endemics [18]. The causative agent of this disease is *Shigella spp*. and mostly *Shigella flexneri* in developing countries [11]. The evolution and the dissemination of multi-drug resistant strains of *S. flexneri* are two of the major concerns in developing countries like Bangladesh, where diarrheal disease rates are the highest, due to socioeconomic and behavioral factors. In our study, we have demonstrated the biochemical and the rapid, specific molecular identification of *Shigella spp*. and *S. flexneri*. The biochemical test TSI revealed that 29 of the 107 non-fermented bacterial cultures isolated by selective plating were *Shigella*. When we performed the molecular test through PCR, we found that all of the bacteria we suspected were *Shigella*. In this work, it was demonstrated that TSI is a reliable biochemical test for *Shigella* detection, allowing diagnosis centers to safely detect *Shigella*. Although the goal of our study was to work with *S. flexneri*, the biochemical test -TSI was not completely successful in detecting specific *S. flexneri*. In that case, 93% of the *S. flexneri* could be detected. However, specific *S. flexneri* detection requires molecular tests. As a result, we observed that molecular identification is the most dependable method for the identification of *S. flexneri*. For molecular identification of Shigellosis, we focused on the multi-copied pathogenic gene-*ipaH*, which is found on both chromosomes and plasmids of *Shigella* and *Enteroinvasive Escherichia coli* (*EIEC*) [1]. Non-temperature regulated IpaH protein, is also distinct from other invasive proteins such as IpaA, IpaB, IpaC, and IpaD both immunologically and at the DNA level. This *ipaH* gene is therefore an excellent target for molecular identification of *Shigella* [1]. We found this gene in all of the isolates. This finding indicates that *ipaH* is an important tool in the identification of shigellosis, allowing us to distinguish *Shigella* from non-fermented bacteria. This molecular method allows diagnosis centers to detect shigellosis with the greatest accuracy. Although this method is somewhat costly, it will assist shigellosis patients in receiving proper treatment. For specific identification of *S. flexneri*, we targeted the antigen specific *rfc* gene, which encodes O-antigen polymerase that polymerizes lipopolysaccharide (LPS) chains. This gene is only found in the genome of *S. flexneri* [10]. Through this gene identification, we found 27 *Shigella flexneri* out of 29 *Shigella* isolates. According to this data, *S. flexneri* was found to be the most responsible agent for Shigellosis in our country. This kind of findings were also reported by P. Parajuli *et al*. in 2019 [11]. According to our findings *S. flexneri* was responsible for 93% (n=27) of shigellosis in Chattogram, Bangladesh. In order to an accurate molecular identification, we have established a positive control by sequencing (*Shigella flexneri*_MZS_191, GenBank accession no: MW774908.1).

After the proper identification of *S. flexneri*, we assessed the prevalence of Shigellosis. According to our investigations, 28 children with shigellosis out of 29 were under the age of five (Table-2). Even more concerning fact is that among these children, 58.62% (n=17) were aged 7 months to 1 year (Table-2). Another interesting finding in our study was that the prevalence of shigellosis as well as other diarrheoal cases are higher in boys than in girls (4A). In addition, the identification and the prevalence of Shigellosis, we also investigated the other causes of diarrhea. Maximum diarrhea cases (47.5%, n=97) were caused due to *E*.*coli/Klebsiella* infection (Table-2). *Salmonella paratyphi* also causes a majority cases (34.8%) of diarrhea. In diarrheoal pediatric patients, *Salmonella typhi, Pseudomonas aeruginosa* were also identified. It is expected that our study will help in rapid and accurate identification of *Shigella spps*. and *S. flexneri* among diarrheoal patients at molecular level that will play role in proper treatment of Shigellosis.

## Data Availability

All relevant data are within the manuscript

## Author Contributions

Conceived and designed the experiments: A M M A C, A K M Z H. Performed the experiments: A K M Z H, M Z H, N S. Analyzed the data: A K M Z H, A M M A C, S A M. Funding acquisition, Project administration: A M M A C. Supervision, resources: A M M A C, Original draft preparation: A M M A, A K M Z H., Review and editing: A M M A C, A K M Z H, S A M, M Z H, N S.

## Funding

The authors received financial support from University Grant Commission, Bangladesh for conducting this research. Fund Reference Number: 37-01-0000-073-04-013/2019/1750

## Acknowledgement

We are thankful to the teachers and staffs of the Department of Genetic Engineering and Biotechnology, University of Chittagong, Bangladesh. Gratitude also goes to Md. Mohabbat Hossain and Pabitra Debnath.

## Declaration of conflicting Interests

The authors declare that they have no conflicts of interest.

## Notes

### Competing Interest Statement

The authors have declared that no competing interests exist

### Author Declarations

“Institutional Review Board”, Chattogram Maa-O-Shishu Hospital Medical College, Chattogram, Bangladesh

